# Where did it go? How Information Management Practices Impact Rigor and Reproducibility in Collaborative Clinical and Translational Research

**DOI:** 10.64898/2026.10.26.26366545

**Authors:** Shruthi Venkatesh, Betsy Rolland

## Abstract

**Introducation:** Clinical and translational research teams (CTRTs) use a vast array of information (digital artifacts used to generate, share, and retrieve data) in their collaborative work, yet information management is often devalued compared to more visible aspects of data-related work. In this second paper from the Information Management Prototype for Clinical and Translational Research (IMPACT-CTR) study, we present (1) the shared practices CTRT members implemented to manage, store, retrieve and share their information, and (2) the impact of these practices on the teams and their work.

**Methods:** We conducted individual semi-structured interviews with 52 team members across 11 CTRTs.

**Results:** We identified information practices that spanned research activities such as team meetings, data collection, dissemination, and overall project management. Effective management of information fostered workflow efficiency, increased shared awareness, and built confidence in the research CTRTs produced. However, inconsistent management of information slowed the workflow, created frustration, and reduced trust in the completeness of research outputs.

**Discussion:** These results demonstrate that information practices are deeply interdependent, preserve institutional knowledge, and ultimately bolster the efficient and effective progression of scientific work. CTRTs who took time to reflect on, implement, and evaluate several lightweight practices through the research lifecycle enhanced the rigor and reproducibility of their science.

## Introduction

> “Different people’s brains work differently. And a [foldering] system that makes perfect sense and seems extremely clear and well organized to one person is just totally opaque to someone else…we haven’t implemented an extremely micromanager-y, impossible-to-follow, one-person’s-brain system, which is good.” (Penelope, Project Manager)

Penelope’s observation captures the personal, and often esoteric, nature of information management. Individuals organize, label, retrieve, and store information to complete their work in ways personalized to their habits and contexts. When groups of individuals come together to work toward a specific goal as a team, these individual information behaviors compound, sometimes clash, and can produce gaps and friction if not communicated properly across members.

Clinical and translational research teams (CTRTs) are unique in their complexity, in that they comprise individuals across diverse roles, disciplines, and competencies [1–3]. Such diversity is embodied by the characterization of “T-shaped translational scientists,” or those who possess both deep disciplinary knowledge as well as the breadth of skills needed to navigate the intricacies of the academic biomedical research ecosystem [4]. These scientists must also demonstrate qualities such as cognitive flexibility and intellectual humility [4] which can help teams build psychologically safe cultures of trust and openness [5]. Collaborative success for CTRTs thus not only depends on alignment across the interdisciplinary diversity of team members, but also on effective coordination across these roles and backgrounds.

The crux of such coordination is information. Information in scientific research is the human-generated digital objects of the work needed to generate, use, manage, and report on data [6] that enables CTRTs to progress along the translational continuum. Indeed, teams are information-processing units [7] and the behaviors around generating, sharing, retrieving, and sense-making of information are integral to the different kinds of work CTRTs engage in. These teams juggle operational work (administrative tasks to support scientific goals), collaborative work (coordinating individual tasks to support the whole), structural work (tasks to establish the collaborative project and meet regulatory requirements), and data work (tasks involved in generating high-quality data) [8]. Despite the central role of information in the functioning of CTRTs across these types of work, there is relatively little literature that delves into the information management practices of these teams.

Our pilot research and initial findings demonstrated that members of CTRTs constantly managed a multitude of information, spread across various tools, often having to adapt and compromise on their individual information styles to align with those of the team’s, to varying degrees of efficiency [9]. They rarely received formal assistance in maintaining their information [6], and piecemeal, individual approaches to managing information led to conflict and slowed scientific progress [10]. Moreover, unclear information management expectations caused frustration and in-the-moment delays in their work, whereas transparency fostered accountability [9,10]. Taken together, our initial findings highlight team members’ need for additional guidance in navigating their team’s information. In this paper, the second in a series from the National Library of Medicine-funded Information Management Prototype for Clinical and Translational Research (IMPACT-CTR) study, we seek to address this gap by identifying information practices CTRTs use to maintain and coordinate their collaborative tasks. We mitigate some of the opaqueness Penelope highlighted by presenting evidence-informed information management practices that have supported team members in completing their scientific tasks and endeavors.

### Key definitions

As described earlier, we define *information* as human-generated digital objects that are used to produce, generate, share, and manage data. Information is socio-technical in nature (lying at the intersection of human-generation and technology [11]) and is myriad in type at each stage of the research process from defining a research question, managing projects to facilitating team meetings. Information is distinct from *data*, defined by the National Institutes of Health as that which can be used to validate and reproduce scientific findings [12]. Data is recognized as the essential foundation for scientific rigor and progress, and much attention has been given to how CTRTs should collect, store, and disseminate data. However, the NIH does not count materials like laboratory notebooks, case report forms, logic models, meeting agendas, or manuscript drafts as data, and there are no requirements to maintain, submit, or standardize these artifacts. These digital artifacts instead constitute information: they provide critical context for the generation and maintenance of data and other project-related tasks CTRTs use in their everyday operation. Each of these artifacts (see [9] for an information typology) either supports the training of personnel to collect data, contributes to data collection and cleaning, facilitates the dissemination of findings, or maintains overall project progress.

Information is likewise distinct from *knowledge*. While knowledge is integrative with information, it extends beyond information: knowledge refers to the productive use of information, as well as the awareness acquired through familiarity, learning, and experience [13], which is beyond the scope of our manuscript.

*Information practices* are ways in which individuals continually “do” information, or the communal ways in which they seek, store, and find information which are specific to a context [14]. In our first paper from the IMPACT-CTR project, we examined how the *information behaviors* (or the individual actions and motivations behind seeking, using, and sharing information) of CTRTs are distributed across a constellation of tools and resources. In this paper, we shift focus to information practices to highlight the habitual ways in which team members manage their information to support their scientific endeavors.

### The Current Study

This manuscript builds on our prior research and aims to identify and share the information practices CTRTs follow to manage, store, organize, and retrieve the large volume of information they engage with to support their scientific, operational, data, and project-related workflows. Having identified key challenges and pain points with information management [6,9,10], we now turn to what works for CTRTs, with a focus on critical tasks members engage in on a regular basis such as managing shared folders, maintaining document versions, and updating participant visit notes. Our analyses of the IMPACT-CTR study follow the Translational Team Science Hierarchy of Needs. This model, proposed in [10], suggests that CTRTs must first meet their basic information and collaborative infrastructure needs before they can cultivate psychological safety and strong team processes to finally achieve translational synergy. In our initial findings, we examined the base of the pyramid, focusing on the collaborative information infrastructure comprising of people, tools, and resources that CTRTs use. In this current paper, we build on our interview data to explore the second layer: the “information management” tier of the pyramid. Specifically, we ask:

RQ1: What information practices do CTRTs implement to support their work?

RQ2: What is the impact of information management practices on CTRTs and their work?

## Methods

Teams were recruited through contacting CTSA Hub admins, word-of-mouth, and internal outreach at the University of Michigan (see [15] for details). The inclusion criteria for the study were that teams had to be U.S.-based funded CTRTs who had at least four team members (inclusive of the PI) interested in participating.

Once a team was enrolled in the study after an informational call with the study team to confirm eligibility, team members were sent a brief enrollment survey (see Figure 1 for study flow). Members then completed a virtual semi-structured individual interview with the first or senior author about their team’s information management practices. The average interview length was 47 minutes.

**Figure 1:**
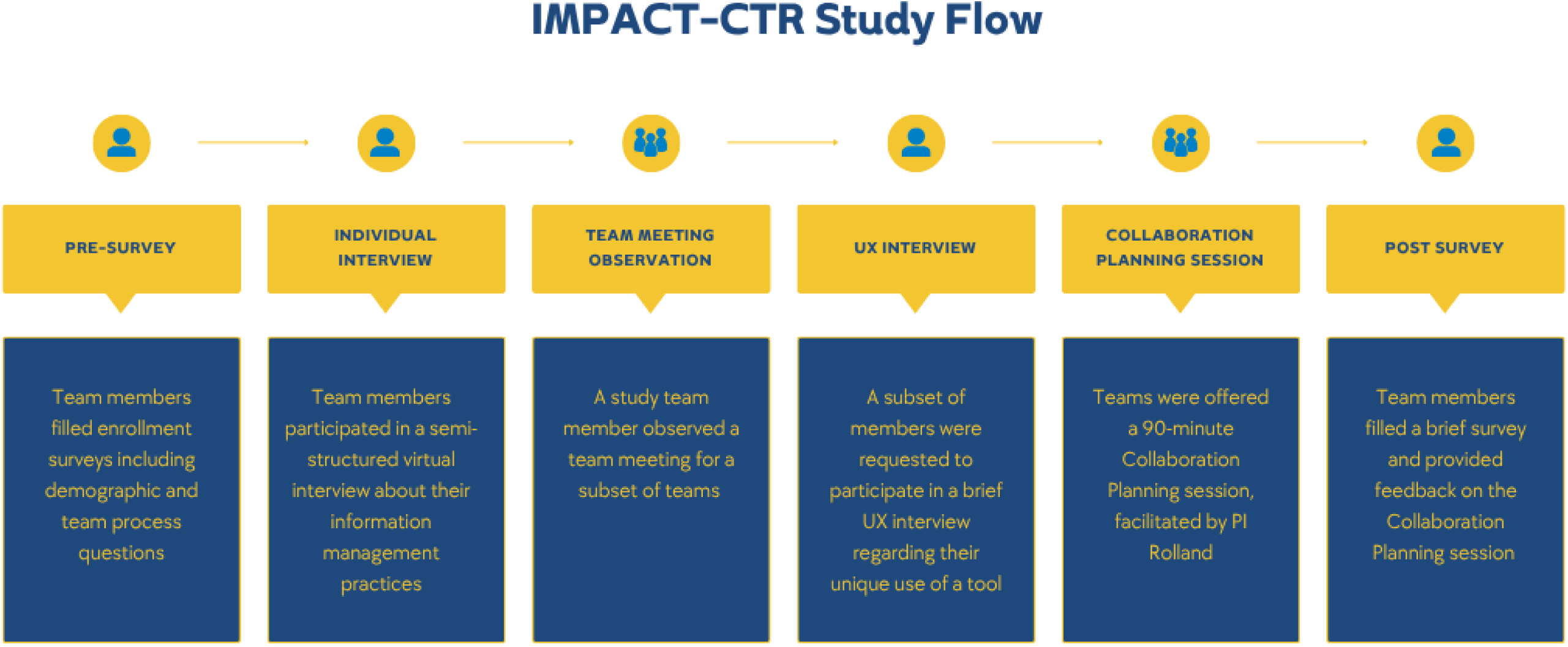
**IMPACT-CTR Study Flow.**

The first author observed a team meeting for a subset of teams and requested to conduct a user interview with some team members who shared a unique use of a tool to manage their information. All teams were offered a 90-minute Collaboration Planning session developed and facilitated by the senior author [16,17] after which participants completed a post-survey with feedback on the session and their team processes. Detailed procedures, the interview protocol, and the data analyses approach have been published previously in [9]. Of note, the first publication reported on 10 teams; in this manuscript, we incorporate data from one additional team. This study was deemed exempt by the University of Michigan’s Institutional Review Board, HUM00255826.

### Participants

Our sample comprised of 52 members across 11 CTRTs. The team size ranged from three to seven members (including the PI), averaging 5 members per team. Thirty-three participants (63%) identified as White, 47 (90%) as Not Hispanic or Latino, and 33 (63%) were female (Table 1). We had 21 Faculty Members including our 11 contact PIs, 17 Staff members, 8 Project Managers and 6 Research Assistants. As described by the PIs, six teams were focused primarily on behavioral and social science, two on biomedical science, two on engineering, and one on clinical science. With respect to geographic distribution, four teams were primarily located in the South, three in the Northeast, three in the Midwest, and one in the West; four teams had members based in different states.

**Table 1:**
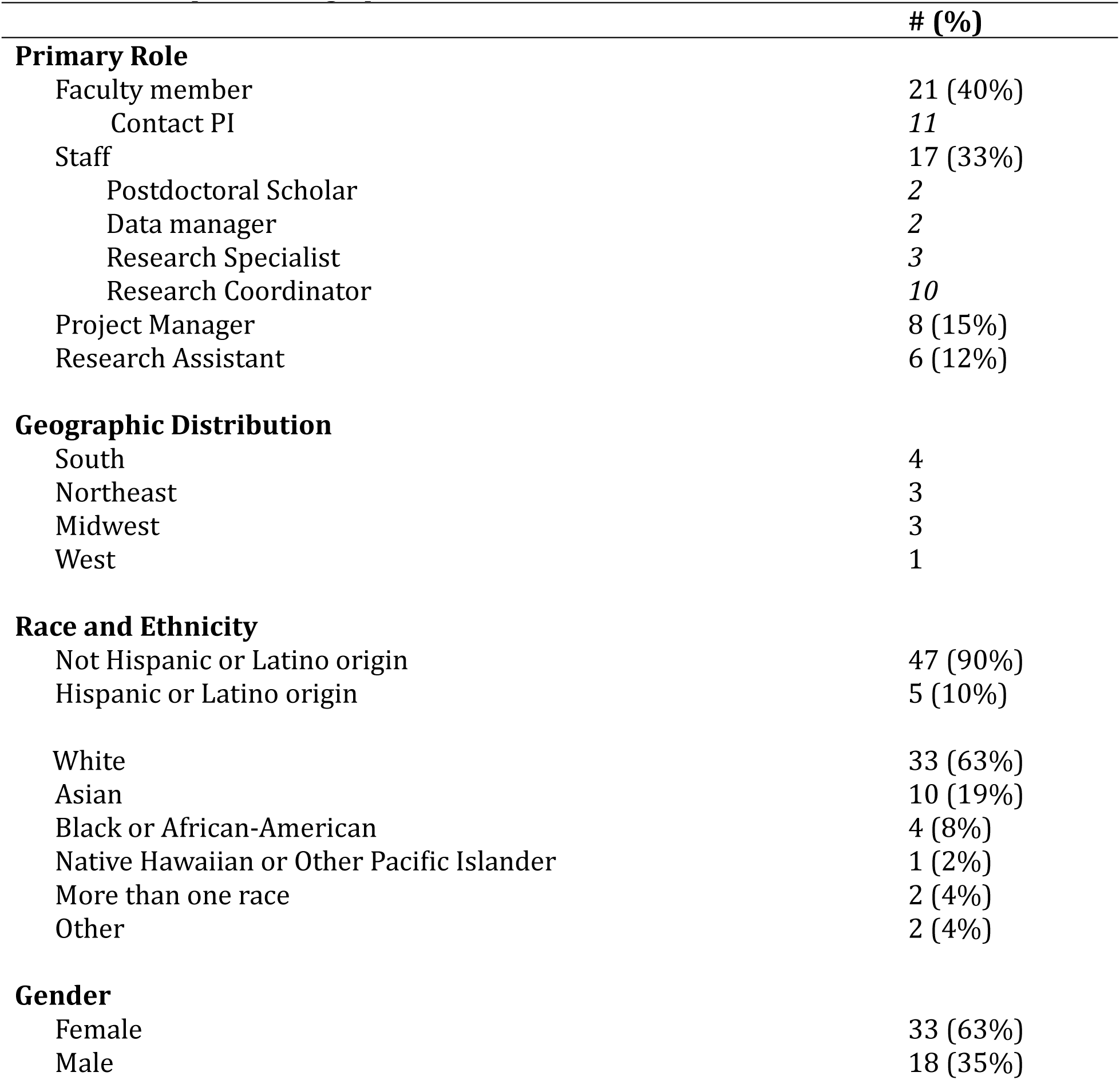
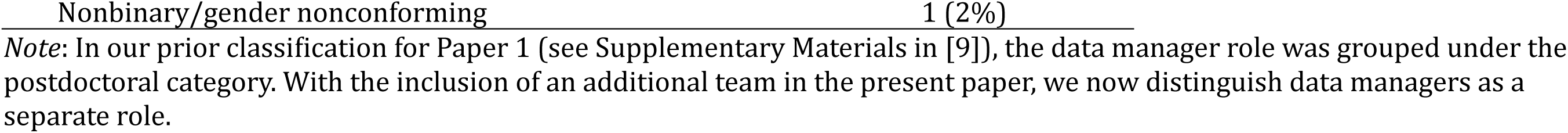
Participant Demographics.

|  | # (%) |
| --- | --- |
| **Primary Role** |  |
| Faculty member | 21 (40%) |
| Contact PI | 11 |
| Staff | 17 (33%) |
| Postdoctoral Scholar | 2 |
| Data manager | 2 |
| Research Specialist | 3 |
| Research Coordinator | 10 |
| Project Manager | 8 (15%) |
| Research Assistant | 6 (12%) |
| **Geographic Distribution** |  |
| South | 4 |
| Northeast | 3 |
| Midwest | 3 |
| West | 1 |
| **Race and Ethnicity** |  |
| Not Hispanic or Latino origin | 47 (90%) |
| Hispanic or Latino origin | 5 (10%) |
| White | 33 (63%) |
| Asian | 10 (19%) |
| Black or African-American | 4 (8%) |
| Native Hawaiian or Other Pacific Islander | 1 (2%) |
| More than one race | 2 (4%) |
| Other | 2 (4%) |
| **Gender** |  |
| Female | 33 (63%) |
| Male | 18 (35%) |

*Note:* In our prior classification for Paper 1 (see Supplementary Materials in [9]), the data manager role was grouped under the postdoctoral category. With the inclusion of an additional team in the present paper, we now distinguish data managers as a separate role.

### Data analysis

In this paper, we focus on data from individual interviews. Deidentified transcripts of interview recordings were analyzed using a hybrid approach, combining inductive and deductive methods [18]. After establishing inter-coder reliability with a research assistant (Kappa = 0.72), the first author independently coded the remaining transcripts. Codes were grouped to pull out emerging themes. Here, we report on the themes pertinent to the research questions for this manuscript.

## Results

### What information practices do CTRTs implement to support their work?

CTRTs engage in research activities across their project’s lifespan, including: (1) holding team meetings, (2) conducting a study, (3) sharing results, and (4) maintaining project management (see Figure 2 in [9]). To complete these research activities, CTRTs implemented information practices designed to help them achieve their scientific goals (Figure 2, Table 2).

**Figure 2:**
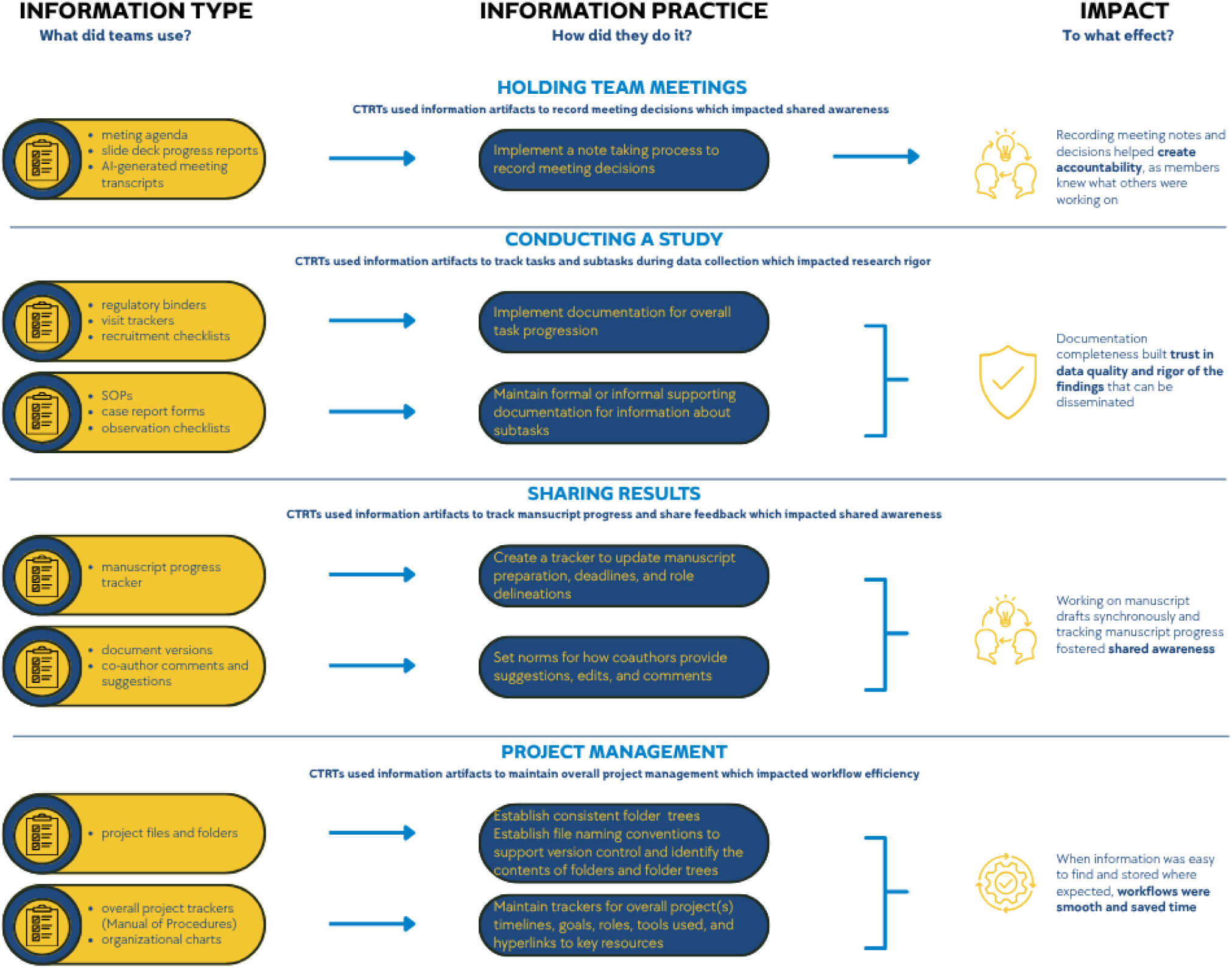
**Information Practices followed by CTRTs,**

**Table 2:**
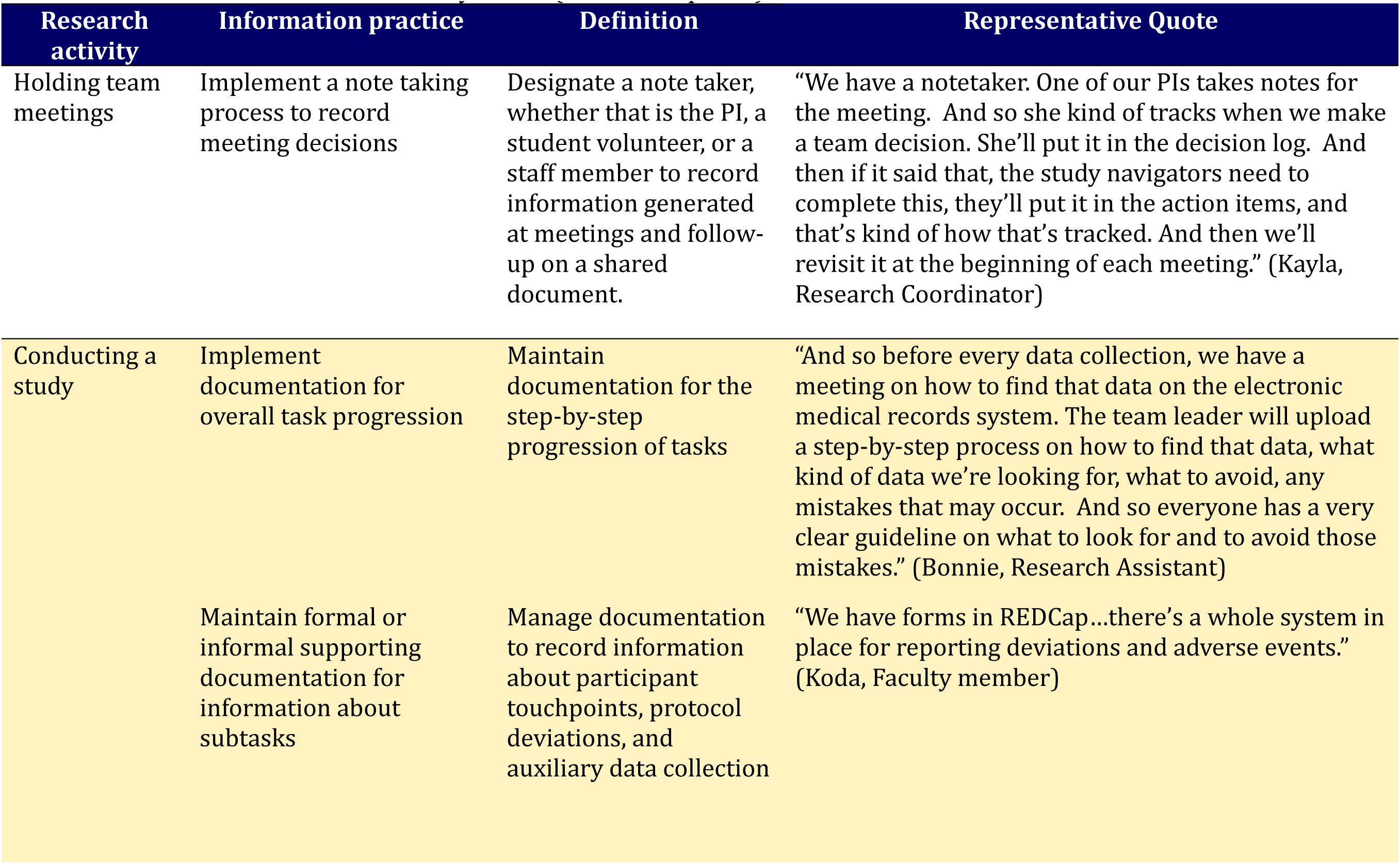
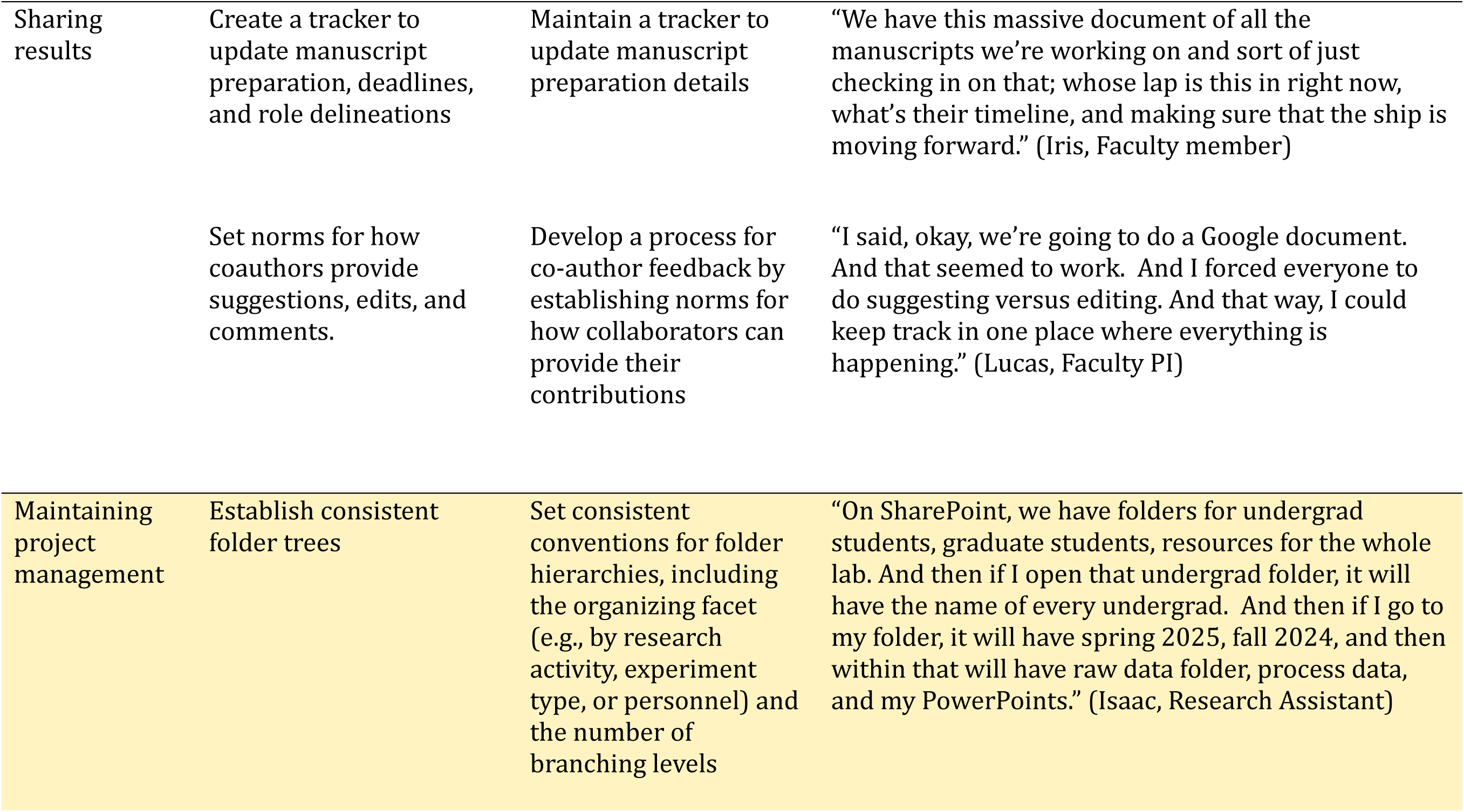
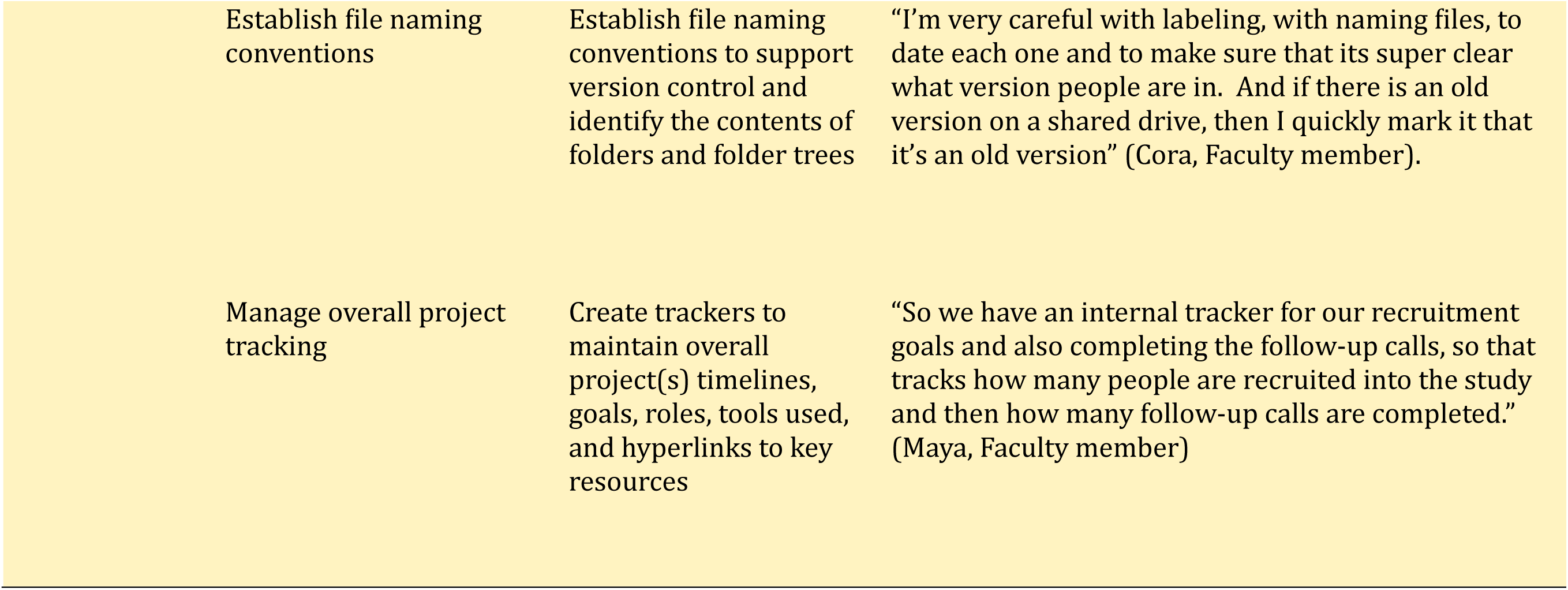
Information Practices followed by CTRTs (additional quotes)

| Research activity | Information practice | Definition | Representative Quote |
| --- | --- | --- | --- |
| Holding team meetings | Implement a note taking process to record meeting decisions | Designate a note taker, whether that is the PI, a student volunteer, or a staff member to record information generated at meetings and follow-up on a shared document. | "We have a notetaker. One of our PIs takes notes for the meeting. And so she kind of tracks when we make a team decision. She'll put it in the decision log. And then if it said that, the study navigators need to complete this, they'll put it in the action items, and that's kind of how that's tracked. And then we'll revisit it at the beginning of each meeting." (Kayla, Research Coordinator) |
| Conducting a study | Implement documentation for overall task progression | Maintain documentation for the step-by-step progression of tasks | "And so before every data collection, we have a meeting on how to find that data on the electronic medical records system. The team leader will upload a step-by-step process on how to find that data, what kind of data we're looking for, what to avoid, any mistakes that may occur. And so everyone has a very clear guideline on what to look for and to avoid those mistakes." (Bonnie, Research Assistant) |
|  | Maintain formal or informal supporting documentation for information about subtasks | Manage documentation to record information about participant touchpoints, protocol deviations, and auxiliary data collection | "We have forms in REDCap...there's a whole system in place for reporting deviations and adverse events." (Koda, Faculty member) |
| Sharing results | Create a tracker to update manuscript preparation, deadlines, and role delineations | Maintain a tracker to update manuscript preparation details | "We have this massive document of all the manuscripts we're working on and sort of just checking in on that; whose lap is this in right now, what's their timeline, and making sure that the ship is moving forward." (Iris, Faculty member) |
|  | Set norms for how coauthors provide suggestions, edits, and comments. | Develop a process for co-author feedback by establishing norms for how collaborators can provide their contributions | "I said, okay, we're going to do a Google document. And that seemed to work. And I forced everyone to do suggesting versus editing. And that way, I could keep track in one place where everything is happening." (Lucas, Faculty PI) |
| Maintaining project management | Establish consistent folder trees | Set consistent conventions for folder hierarchies, including the organizing facet (e.g., by research activity, experiment type, or personnel) and the number of branching levels | "On SharePoint, we have folders for undergrad students, graduate students, resources for the whole lab. And then if I open that undergrad folder, it will have the name of every undergrad. And then if I go to my folder, it will have spring 2025, fall 2024, and then within that will have raw data folder, process data, and my PowerPoints." (Isaac, Research Assistant) |

### Research Activity 1: Holding team meetings

All teams in our sample held team meetings at a regular cadence. Information generated at meetings included progress updates, action items, to-dos, decisions, and task assignments. We use the term “decisions” to encompass information that clarified responsibilities, procedures, or other directions needed for team members to move their projects forward.

#### Information practice 1: Implement a note taking process to record meeting decisions

An information practice followed by most teams was to implement a note taking process to record meeting decisions. This practice typically involved assigning a designated note taker, whether the PI, a student who volunteered, or a staff member, to ensure someone was responsible for capturing the information and decisions generated at these meetings. Some teams also had the AI summary feature enabled for virtual meetings.

Without a designated note-taker, there could be lapses in recording meeting decisions, as one member noted that there was no formal appointment in their team: if their volunteer note-taker did not attend the meeting, the consistency of their note-taking would be negatively affected.

Majority of our teams had a shared living document for meeting agendas and notes that resided on the team’s shared storage. This document could also include links to AI summaries or AI transcripts, if those features were enabled. Having such a shared digital artifact served as something “durable” that the team could refer to at any point. Indeed, this shared meeting document was seen as a go-to resource by many members if they needed to revisit a decision that was made collectively during a meeting. Additionally, a couple PIs mentioned that they referred to these meeting minutes when gathering information to write a manuscript, “If I needed to confirm how something was done, I would go back to our meeting minutes and look for where it was discussed” (Mabel, Faculty PI).

Typically, the person assigned to take notes also handled post-meeting follow-up, which included either tagging members responsible for specific tasks in the agenda document or circulating an email with the list of tasks to be done. As a few team members noted, team meetings served as a space to gather information on “who needs help and who wants to give help.” Iris elaborated on how this information was shared during the meeting,

> “So the main thing was making sure that if someone was out for a little bit, that someone else was covering their timepoint. And so we would just, in the minutes, collect ‘I’m still doing the six-month calls’. And then if, ahead of time, they all knew that if they can’t make a meeting, they have to give us an email update ahead of time. So then we would just add that to the minutes. So-and-so is still working on this. They just can’t make it today. And then if someone can’t do it, then we would just email that person and say, ‘hey, can you take over this person’s calls.’” (Iris, Faculty member)

### Research Activity 2: Conducting a study

Two practices emerged relating to maintaining, using, sharing, and storing information about study data: (1) Implement documentation for overall task progression, and (2) Maintain formal or informal supporting documentation for information about subtasks. To note here, we use the term *documentation* to refer to artifacts created in any digital format, whether they are created in the Microsoft Suite, Google Docs, spreadsheets or in project management tools such as Asana. Similarly, *information about data* refers to artifacts such as laboratory notebooks, case report forms, notes about protocol deviations, observational notes during participant visits and so forth that is not “data” as defined by the NIH, but integral to the production, usage, and subsequent understanding of the data generated. Finally, we use *task* to refer to the higher-level jobs to be done, and *subtask* to refer to the smaller steps within those tasks.

#### Information practice 2: Implement documentation for overall task progression

Research tasks are often complex, requiring multiple steps to complete. An information practice that enhanced task completion was having step-by-step documentation of overall processes that a CTRT needed to follow to collect required data.

Team members developed their own versions of checklists, such as written guides, visit trackers, regulatory binders, and spreadsheets to aid in the sequential progression of these tasks. Although these trackers varied in form, the underlying approach was the same: having a defined path from steps A to B to C. For example, Benjamin, a Project Manager, described the overall process for data collection that the research assistants on his team followed,

> “We created rules on who documents what. [Research assistant] recruits. Then they send us [clinical center] this email. That’s their documentation, and we respond to that. Then when they [research assistants and participant] come in-person, they bring the consents with them. The consent process happens. It is documented, and then we [clinical assistants] do our visit, and we document again that the visit has been completed in Epic. We fill out the case report forms. We upload the consent in Epic, so then, the patient has the consent uploaded into Epic for themselves. So we kind of created who does what.”

Such process checklists lived on shared storage and were used as a training guide by some teams for new members. These overall task progression trackers were especially useful where the responsibilities of recruiting, collecting data form participants (or animal models), and analyzing the data were delegated among members.

#### Information practice 3: Maintain formal or informal supporting documentation for information about subtasks

In addition to maintaining documentation about the overall sequential tasks involved in conducting a study, CTRTs developed supporting documentation for the various steps within those tasks. The Standard Operating Procedures (SOP) was the most common type of checklist, typically mandated by the IRB, and had to be followed for certain standardized subtasks such as participant observation, blood draws, or processing experimental reagents.

Apart from such formalized documentation, team members had also created informal documentation to capture information about the data collection process that was not always required by the SOP, but instrumental for the team’s records of what had happened during data collection. For instance, referring to Benjamin’s example mentioned earlier, while the IRB included the tasks of recruitment and bio sample collection, each of those tasks required a number of subtasks to be defined, described, and tracked. For instance, a research coordinator on the same team shared how they maintained a tracker to log every contact with a participant. Such practice was common,

> “I had them [students] track by the activities that they did with participants in the REDCap database. So any time they had an interaction with the participant, because it was longitudinal, they documented what they did with them. So they can go back there and see. So that was part of the protocol, so that we did have documentation all throughout the process, and it wasn’t just like willy-nilly, where we get stuff from.” (Shane, Faculty PI)

The entries on such trackers would not constitute data per se that the team would eventually report on. However, the existence of such artifacts facilitated shared awareness by helping team members see what others had done, since data collection was rarely conducted by a single individual and often required coordination around where work had been left off and what had been completed,

> “The recruitment tracker actually helps, you’re like, ‘I don’t know what’s next. Let me see.’ But if you have it right there in black and white, you can kind of look at it…you’re not duplicating. I can see [team member] tried to reach [participant]15 minutes ago. She sent a text. She left a voicemail. So I don’t want to just start calling [participant], because [team member] already did it.” (Callie, Research Coordinator)

Herein, it was not only important to create multiple checklists or SOPs, but to also ensure team members were implementing them consistently. Roy, a Project Manager, aptly reflected about this when he shared his team’s (lack of) use of inventory logs when setting up the inventory for a data collection visit,

> “The SharePoint link to the inventory log was sent out via email. The issue is, it only works if you do it. You could go to the basement and take a whole bunch of stuff out, but if you don’t update the inventory, then it serves no purpose. If you don’t sign the key out, then the key is still gone, and there’s no oversight.”

Majority of our team members reported knowing about and using these trackers as part of their training and everyday data collection. They could see each other’s regular updates on these trackers that were stored on shared storage. One team even described incorporating attention-check prompts, where members had to comment on their favorite snack on a spreadsheet row to verify that had been following the information flow. For a couple of teams, however, this remained an outstanding issue as Roy alluded: while the team had task trackers, consistent use of them did not always occur.

The details of what occurred during data collection, as documented in these trackers and observational notes, ultimately informed the Methods and Results sections of academic manuscripts. Faculty members described referring to these records when gathering information for manuscript preparation. Kevin, a Faculty PI, noted that the “biggest thing will be those observational notes, making sure we know exactly what was done… And it relies on that each person took careful notes of those things in the first place.” He added that while IRB protocols or grant applications served as a base for information, what was planned does not always reflect what actually happened during data collection. These supporting documents thus provided ways to capture protocol deviations and other aberrations that may occur. Hence, maintaining complete trackers and checklists supported the conduct of rigorous, reproducible research and also facilitated subsequent manuscript preparation.

### Research Activity 3: Sharing results

More than half of the members in our sample contributed to manuscript preparation and publication. Manuscripts were typically led by PIs or faculty members, with other team members contributing to various stages of the writing process. Participants described gathering the information needed for manuscript preparation from previously published protocol papers produced by their lab, IRB protocols, grant applications, and notes documented in the data collection trackers described earlier. Two practices were helpful in streamlining the manuscript preparation process: (1) Create a tracker to update manuscript preparation, deadlines, and role delineation, and (2) Set norms for how coauthors provide suggestions, edits, and comments.

### Information practice 4: Create a tracker to update manuscript preparation, deadlines, and role delineation

A couple of teams described maintaining a manuscript progress tracker, typically a shared spreadsheet that provided a snapshot of all manuscripts that were in progress, recently submitted, or in the ideation phase. The tracker included information such as target journals, submission deadlines, manuscript leads, and the status of individual writing tasks,

> “[We have] a manuscript tracker. So divided up by different areas, we have the manuscript title, data source, authorship, who our community partners are, the status, the last time the status was updated. Who’s in charge of it right now? When are we hoping to submit? What journal? The manuscript [versions] are also hyperlinked on the spreadsheet” (Demi, Faculty PI)

Such a tracker helped the team monitor progress of its scientific output and also promoted role clarity. Team members with access to such a tracker reported having a clear understanding of their writing responsibilities,

> “I’ve had a pretty clear role on these papers. I’ve known exactly what I was supposed to do with the papers, write the background section, add citations in, write these vignettes…very discrete roles, which has been, that’s been easy.” (Faith, Postdoctoral scholar)

The tracker was reviewed periodically during team meetings, which enabled team members to step in and reassign writing tasks as individual availability waxed and waned throughout the semester.

### Information practice 5: Set norms for how coauthors provide suggestions, edits, and comments

CTRTs followed a variety of collaborative writing processes during manuscript preparation. Irrespective of the process, typically developed through trial and error by the PI’s experiences, one way to maintain the information generated during manuscript preparation (such as document versions, comments, figures, supplementary materials) was to establish an explicit process for how, where, and when co-authors should contribute.

Establishing and communicating the preferred process of information flow from coauthors during manuscript preparation helped reduce the chances of overwriting. Some teams had moved away from Word documents, citing the difficulty of integrating multiple tracked changes versions, and instead had adopted Google Docs. Here, they enabled the “suggesting” mode that served as a synchronous way for authors to review and integrate feedback. Other teams relied on a sequential writing process, in which the lead author(s) drafted an initial version of the manuscript and then circulated to their coauthors for specific feedback. A few teams had adopted more visual approaches for idea generation. For example, one team used Mural boards (digital visual workspaces) to brainstorm points for a Discussion section. The lead author then used affinity mapping to cluster ideas and draft the text. Another team reported using whiteboards,

> “And then he [PI] documented his thinking, like a flowchart type… I would say he has a whiteboard in his room, and we did a lot of in the moment, just webs and diagrams, and translated that onto the paper and then onto email.” (Isaac, Research Assistant)

Coupled with the manuscript tracker, these practices also conveyed that “there is enough work to go around and we can all have a piece of the pie” (Nyla, Faculty member).

### Research Activity 4.1: Maintaining project management: Folder structures

All CTRTs in our sample used shared storage platforms such as Box, OneDrive, SharePoint, or Google Drive to host, share, and store both project-related information, information about the data, and the data itself. Organizing, categorizing, and maintaining a consistent structure of these artifacts is one aspect of overall project management.

Information and data were commonly organized using folder trees (i.e. branching structures), with main folders containing nested folders within which files (documents, spreadsheets, PDFs, presentations, graphics, etc.) were housed. However, folder practices exhibited a large variation across the teams in our sample. Some teams reported having folder tree norms, but even within those teams, a couple members described varying levels of initial overwhelm when orienting themselves to the folder structure. Moreover, for the teams who had easier-to-navigate folder systems, it had required individual members to work through the “me vs. we” [9] conundrum of adapting their own organization styles to align with those of their team’s. Other teams did not have established norms such that members determined where to store their files on the fly. This lack of norms often led to additional time spent searching for and locating needed files, “I think most of the time I end up finding it [a file], but it’s taken a while and I’ve lost a significant amount of time. I feel like that’s probably the most common outcome” (Katherine, Faculty member). To ease the difficulty of navigating folder trees, two practices emerged as either helpful to follow or practices that team members wished they had implemented: (1) Establish consistent folder trees, and (2) Establish file naming conventions.

#### Information practice 6: Establish consistent folder trees

CTRTs reported setting (or wanted to set) consistent conventions for folder hierarchy. Characteristics of easier-to-navigate folder structures included that they (a) were consistently “chunked in broad stroke categories” (Derek, Project Manager) such as by experiment date, personnel name, or project task based on how the original owner of the files had structured them (typically the PI or senior project manager), (b) were not more than three layers deep “so you don’t have to go that far down to find what you’re looking for, which I like a lot better” (Shane, Faculty PI), (c) had separate subfolders for archived files that were “accessible” but not “cluttering” the current files (Sophia, Project Manager), and (d) were explained to other team members, by giving new members a “tour” of their OneDrive folder during orientation. For example,

> “For this collaborative effort, there is a Box folder. Within that Box folder, there is a folder for each grant that’s funded. For example, this work is all being done on this [funding mechanism], so now any data files go in that folder. Within the main folder, there’s a separate folder for grants submitted, right? And then within that folder are the working documents for every grant that we’ve submitted, whether or not it was funded. But once a grant gets funded, we make a folder within the main folder for that specific grant, and then any data that is generated for that grant goes in that Box folder.” (Bailey, Faculty PI)

For teams that maintained physical regulatory binders, another helpful practice was to reflect the organization of the physical binder and its documents in the digital shared storage folder, “You build your OneDrive folder basically to mirror a regulatory binder, so you have different sections for different components of the study, you have a regulatory folder and a subject visit folder and a folder to store communication. That’s kind of our go-to system, and it works well” (Daphne, Research Coordinator).

#### Information practice 7: Establish file naming conventions

A complementary practice to consistent folder structures was establishing file naming conventions that specify how both folders and the files within them should be named. Many team members in our sample expressed a desire for clearer naming conventions within their teams, often in hindsight as they reflected on the file organization during their interviews with us, “I guess we could work at thinking about the titles of the folders. We could consider better labeling. You know, what works for one person is maybe not so straightforward for somebody else” (Nyla, Faculty member). Members had experienced trouble locating files because it was not immediately obvious what belonged in a particular folder, “The idea is, with the naming, that it should be obvious by the folder titles what belongs there. And it wasn’t at the time” (Demi, Faculty PI). Additionally, struggling to find documents by their right names was not only frustrating, but also took time away from task progression,

> “There can be some confusion where somebody looks at, say, Figure 1, and then they’re trying to find the source code, but they’re looking at the source code for Figure 2, and it doesn’t quite match. Obviously, they’re not the same thing. And so that can require a little bit of troubleshooting and man hours to finally realize that, ‘oh, this isn’t the code that I want to be running.’ For whatever reason, I looked in the wrong place, or maybe, frankly, maybe it was mislabeled or labeled a little bit ambiguously.” (Derek, Project Manager)

However, creating naming conventions was described as “low priority” or something most teams had not got around to establishing.

In the absence of a formal naming convention, individual members shared their workarounds, such as, (a) giving files descriptive names so they would appear using search functions, “I give my files very descriptive names. And so as long as I could get at least one of those words right, it’s going to come up, you know?” (Chloe, Faculty PI); and (b) naming files with dates on them, “Just give everything a date. Just be safe” (Caleb, Research Assistant). Nancy, a Research Specialist, reported on her team’s work-in-progress with respect to naming their experimental particles. Her team, which collaborated across five labs, had had inconsistent naming practices. She had been “instrumental” in creating a naming convention, so they now followed a naming system that was managed via an overall experimental log. It was partially retroactive, wherein she had renamed particles analyzed previously with the aim of preventing naming inconsistencies from percolating into future work,

> “We changed the way that we call our particle. And at one moment it’s ‘how we compare this name now to the name that we used in the first experiments a year and a half ago?’ And sometimes it’s not clear, but we decide, from now, and we put the note, from now, we call this one that was referred to that one in that experiment. And maybe it’s not sufficient, but now I know what you are talking about.”

Another benefit of establishing naming conventions was to improve version control management. It was only natural for CTRTs to have multiple versions of files as they drafted manuscripts, submitted amendments, or underwent study protocol changes. Handling multiple versions of these digital artifacts could become confusing. Hannah, a Data Manager recalled creating copies of codes across statistical platforms like SPSS and STATA, and the challenges of maintaining the most updated version, “So it’s either a matter of, rewrite the code to do the same things in those programs, or just copy and paste in the new variable from one source and just copy and paste it into the other one. You know, just trying to manage things across multiple platforms was a pain.”

Tracking version history through clear labeling of documents and maintaining a distinct separation of current versus old or archived files helped make clear what the most current version of a document was. For example, Diana shared her team’s approach to copying and labeling manuscript drafts,

> “So even with manuscripts, what he [PI] does is the first draft is V1. And let’s say everyone made comments on V1, so what he would have us do is make a copy of it, and that’s V2. And that’s where you address all the comments. So even if it’s not currently relevant, I can always go back and find it.” (Diana, Research Assistant)

### Research Activity 4.2: Maintaining project management: Overall trackers

Some CTRTs also maintained overall project management trackers that served as a high-level overview of the team and its ongoing project(s).

#### Information practice 8: Manage overall project tracking

Project trackers took on a couple different formats. One format, whether in the form of shared spreadsheets or on tools such as Asana, provided a big-picture preview of the team’s projects, people, protocols, and software tools with relevant hyperlinks to how-tos and team processes. In [9] we refer to the “Super Spreadsheet” as an example of a spreadsheet that housed this information. For instance, Sophia, a Project Manager described how this spreadsheet contained rows for each type of research task that was currently active, with columns for which undergraduates were assigned to the task, deadlines, and links to supporting files.

Other formats supported more specific aspects of project management, such as Gantt charts for tracking timelines, or organizational charts for documenting team structure. To illustrate, Isaac described how it was beneficial to have an organizational chart that delineated who he reported to within the lab’s hierarchical structure, “PI has a chart of our entire lab and the organization, so like project one, two, three, graduate student one, graduate student two, and the associated undergrads beneath them. And so that gives a very clear image of where do I fit in in the lab. And that makes it really clear for me, who do I need to go to” (Isaac, Research Assistant).

Independent of the format, these project trackers could also serve as guides to orient new team members to the team, personnel, and project goals. For instance, Nyla reflected how their team did not currently have such a tracker though it would be a nice-to-have,

> “And so it might be nice to have an introduction document to the team, like what we are working on, what our goals are, and a running list of team members. And then in that same document, you could have just reminder points about how we save data, how we share data, who to reach out for help. It might be nice to have one document that kind of is something that you can refer back to describe the team process and protocols and people.” (Nyla, Faculty member)

### What is the impact of information management practices for CTRTs and their work?

The aforementioned information practices supported the different types of research activities CTRTs engaged in. Three overarching impacts, or the possible costs and benefits of following (or not following) these practices emerged from the data: (1) workflow efficiency, (2) shared awareness, and (3) research rigor. Ultimately, while each practice affected all three impacts in some way, particular practices had a more profound impact on certain outcomes than others.

### Workflow efficiency

Workflow efficiency refers to how an information management practice affected the speed and ease of the tasks CTRT members had to carry out. Information was perceived as being straightforward to manage, use, or store when members knew where to locate it and could access it easily and quickly.

Information practices relating to project management had an especially strong impact on workflow efficiency. When files were easily found and/or located in the folders they were expected to be in, it fostered smooth workflows,

> “It [work] goes a lot smoother because, for example, I was able to go into that [community collaborative] folder and easily be able to pick out what I needed because the Manual of Procedures said directly, when you’re done video recording community collaborative, this is where the notes go. And I was able to just follow that, go into the folder, get the notes that I needed, and implement the feedback” (Diana, Research Assistant)

Whereas, when information was not easily retrievable on shared storage due to confusing folder trees or inconsistent file names, it created frustration, delays, and duplicated work. For instance, Eva described the time lost to searching through folders for a particular file, “Well, just a little frustrating because I know we have it. Where is it? I’ve written this before. It kind of slows the process and adds frustration” (Eva, Faculty member). Similarly, Mabel described having to redo a table because she could not find the original file on Google Drive, “I spent time yesterday recreating the whole table…that was frustrating because it cost me an extra, I don’t know, 45 minutes that I had to do” (Faculty PI). Thus, team members spent additional time tracking down information, reconciling document versions, or even duplicating efforts because they could not locate where a file, table, or code had been stored.

### Shared awareness

This impact moves beyond workflow and into team climate, highlighting how information management practices can affect shared awareness, or the extent to which individual members felt they knew and understood what their other peers were working on, as well as how their own roles contributed to the collaborative effort. Such awareness promoted accountability because due dates were documented on shared documents or team members notated their progress on subtasks such that others could see and know what they were working on. Moreover, shared awareness also fostered transparency by making members’ contributions visible to the rest of the team. For example, having access to one another’s folders or viewing contributions on drafts and spreadsheets gave clarity on what each other was working on, “Folks know what to expect. They know how to keep up with their [recruitment] numbers and gives us a better sense of the progress we are making” (Izzy, Faculty member).

Shared meeting agendas impacted shared awareness as these agendas helped create accountability: members knew what others were working on, when their check-ins were due, and even found it “illuminating” to highlight what had not yet been completed,

> “Linking the notes and the activities within those standing meeting folders has really helped with continuity of communication and making sure things aren’t dropped so I think it’s helped people feel sort of accountable for their task because they know when their check-ins are due.” (Demi, Faculty PI)

In a couple of teams, members reported not having a shared meeting document per se. A few members shared how this lack of a shared document created information gaps: Josie, a Research Coordinator felt he did not have a broader sense of what the rest of his team was doing,

> “It [meeting process] can improve by having links to whatever the important decisions we’re taking for the month or weekly. I need to shadow for different projects because I will be doing some of them. And I think this [documenting] will help knowing what days they’re [other members] going out in the community or just what they’re up to. And learn from them…I think you have a better opportunity just to connect better with the team members and form a stronger bond.”

Maintaining manuscript trackers and working on manuscript drafts synchronously also fostered shared awareness, as Daisy noted, “I like to maintain the single shared version in one place, and then people can jump in and out of it. You can see when other people are in it. And you always make sure that you have the very most up-to-date version… This is where we are. And we can both [staff member] pull it up at any point and see where things are. So we’re actively sharing a live document” (Faculty member). As such, being able to see what other team members were working on, especially for shared tasks and collaborative writing, promoted transparency.

### Research rigor

The information practices around conducting a study directly impacted the scientific rigor of CTRTs’ work. Information management ultimately impacted the effectiveness, quality, and rigor of the data produced. When SOPs were consistently implemented and supporting documentation was maintained, it built confidence and trust in the completeness of information about the data collected, and thus, the data themselves,

> “So [LAB MANAGER] monitors everything she does. She’s got those sheets in OneDrive that every day, she goes to the mouse room, and she takes all her notes. She goes back to her computer and updates everything. And then it’s all stored, and she’s got tabs by week. So I basically could retrieve in an instant every single thing she’s done in the mouse room for 15 minutes. I know where it is, and I know she is so exquisitely detailed about her note keeping.” (Daisy, Faculty member)

Whereas, when such information was managed ineffectively, it led to missing information about protocols, deviations, or how the data were collected, which reduced trust in the rigor of the findings,

> “I discovered that no physical, no notes were ever taken on the symptoms. So other than my meeting notes, I have no record of the toxicity. I don’t know exactly what symptoms they showed. And so as a result…we could have published a study with much more detail, but instead we can basically just comment qualitatively on some observations and on our decision-making. It limited what we could conclude by the fact that we didn’t have the records.” (Kevin, Faculty PI)

Repeated failures to document information about data compromised overall data accuracy and integrity, “We always end up confused about where we’re trying to go. We do experiments that aren’t documented well. I spent probably ten times more time than I typically do to get the information, to get it polished, to fix errors…I think pretty profound challenges in the movement of our science comes about because of this” (Lucas, Faculty PI). This lack of data integrity hindered the team’s scientific progress.

## Discussion

CTRTs’ work is multilayered and complex and spans a variety of research activities simultaneously. Here, we highlight the information practices that were common and useful across the 11 teams in our sample. Our teams used these practices to varying levels of consistency and maintenance. Effective management of information fostered workflow efficiency, increased shared awareness, and built confidence in the research CTRTs produced. However, inconsistent management of information slowed the workflow, created negative emotional experiences, and reduced trust in the completeness of research outputs.

Although we presented the practices in a list-like form, with practices somewhat siloed into specific research activities, in actuality, these practices are highly interdependent. Members of CTRTs work on different tasks at the same time and rely on each other to move their tasks forward [19]. For instance, a research assistant may need to accurately locate the most *current version of a manuscript* before entering her contribution to the paper, including *writing up any protocol deviations* that occurred during data collection. Doing so may require her to trace those deviations back to the *project trackers*, where experiments were documented as they were conducted, potentially by another team member. In this way, members often need to switch across different types of information and ways of managing that information simultaneously to facilitate those shared mental models which are a key aspect for cultivating psychological safety and knowledge sharing in teams [20].

These information management practices also support information continuity and preservation of institutional knowledge. Not only are members of science teams highly interdependent on each other, but their membership on these teams is permeable and fluid [21]. The cumulative effect of preserving the history of key project decisions and maintaining documentation can collectively ensure that tacit learnings are conserved through the creation of digital artifacts that can be used by other team members. These checklists and cheat sheets provided a practical way for team members to document, as they went, what had worked, what had not worked, and what had changed during the conduct of the study and related project management. As one faculty member noted, “I think it’s worth all the time and effort to put all that kind of organization at the beginning because then, people can come and go, but it’s still easy to find stuff, right?”

While these information practices are lightweight to implement, they tend to be devalued in the pursuit of CTRTs’ data and scientific work. The everyday logistics of maintaining and coordinating projects at scale, with team members spread across labs, departments, and institutions can get waylaid. Indeed, publicly funded PIs are expected to not only exhibit research leadership on their projects but also be at the helm of their administrative coordination with little institutional support or training [22]. But this overlooking comes at a cost, as we saw in the impact of inconsistent information practices on reduced workflow efficiency, and, ultimately, the rigor of the scientific findings that these teams are formed to create in the first place.

It is not only important to establish checklists, trackers, norms, and rules at the beginning of a project, but these information practices are only useful insofar as they are implemented and consistently followed. Just as teams take time to discuss raw data, in-progress data, manuscript versions, and other activities that conventionally move their science forward, we purport that they should also collectively make time to discuss and assess the day-to-day information infrastructure that supports these activities. Being proactive about this infrastructure may ultimately enable teams to better pursue their scientific endeavors. More than an evaluative approach, our findings are intended as a way for teams to learn from one another. Our analysis is shared through a lens of understanding, as echoed by some of our participants: science is hard, requires members to often pivot, and there are people behind these complex information layers, “Our systems are not perfect, but we do our best. We really do.” (Fiona, Research Coordinator).

To this end, we are working on translating the findings presented in this manuscript into a lightweight workbook for CTRTs. The goal is to provide a simple, adaptable starting framework that teams can use to reflect on, implement, and evaluate their information management practices through the lifecycle of their projects. Rather than prescribing a standardized approach, we intend that the workbook will aid teams identify and implement practices that align with their own collaborative needs and resources.

### Limitations

One limitation in our analyses is that the practices listed here represent a summative analysis across participants and teams. Scientific teams differ considerably in how they are structured, who they are composed of, and the institutional constraints and resources to which they have access. Teams generally found what worked for them given the tools, the people they had, and the ways in which their teams were structured. As such, some of these nuances were difficult to capture in an analysis such as this, especially given the need to maintain de-identified participant information without revealing details about individual teams.

Another limitation is that the CTRTs in our sample may not be representative of translational research teams more broadly, given the differences in team structure and institutions mentioned earlier. Although we had 11 teams, the findings may not capture information practices used more widely across CTRTs. Furthermore, we draw from individual interviews which only reflect the perspectives of those team members who chose to participate in our study.

### Future Work

In this iteration of the analysis, we remained tool agnostic. As described in our first paper, tools can vary considerably depending on institutional access and come with challenges such as interoperability, baseline tool readiness, and familiarity [9]. Understanding the relationship between information practices and the tools used to support them is an important area for future work, including examining user experience surrounding these tools. For instance, it is conceivable that a team which uses a centralized project management tool such as Asana to maintain its trackers, supporting documentation, and meeting minutes may experience information management differently from a team whose corresponding information is dispersed across Google Drive and Dropbox.

Another direction for future study is to further examine the relationship between information management practices and team outcomes. While we identified information management practices and observed their potential effects on team functioning, these were primarily gleaned by asking participants about how they experienced the impact of these practices. Thus, additional research is needed to effectively measure which practices have the greatest impact, how these practices change over time, and what results from implementing or not implementing them. Development of such measures will also add to broader conversations in the team science literature on the evaluation of team outcomes [23].

### Conclusion

Taken together, these findings highlight the integral role of information practices in supporting the collaborative work of CTRTs. From documenting meeting decisions to coordinating overall project activities, these information practices help manage the complex and layered types of information team members navigate in the daily conduct of their tasks. The lightweight, accessible practices presented here require modest effort to implement and are essential to collaborative science. However, teams struggle to find the time, resources, capacity, or frameworks to implement and integrate these practices through their project life cycles. We aim to provide CTRTs with these adaptable evidence-informed practices to strengthen their information management, and ultimately support more effective, efficient, and rigorous science.

## Data Availability

All data produced in the present study are available upon reasonable request to the authors

## Acknowledgements

We thank Savitha Sangameswaran for her help with creating the data collection methods, and Jamil Rkein for his support with coding. We thank the team members for their participation in our study.

